# Juvenile and adult open-angle glaucoma form a single polygenic continuum

**DOI:** 10.64898/2026.09.13.26361381

**Authors:** Joshua Schmidt, Ma. Carmela Guevarra, Guiyan Ni, Thomas A. D. Hassall, Shannon Ji, Thi T. Nguyen, Georgina L. Hollitt, Antonia Kolovos, Emmanuelle Souzeau, Ngoc Quynh Le, Puya Gharahkhani, Jason Turner-Maier, Edward Ryan A. Collantes, Inas F. Aboobakar, Ayellet V. Segrè, Alex W. Hewitt, Nick Haan, Stuart MacGregor, Janey L. Wiggs, Jamie E. Craig, Owen M. Siggs

## Abstract

**Purpose:** To test whether juvenile open-angle glaucoma (JOAG) and primary congenital glaucoma (PCG) are polygenically continuous with primary open-angle glaucoma (POAG), using a multi-trait POAG polygenic risk score (PRS).

**Design:** Retrospective cohort study.

**Participants:** 3,102 open-angle glaucoma index cases, of whom 2,836 had a recorded age at diagnosis (52 PCG, 226 JOAG, and 2,558 POAG) and 999 European-ancestry controls from a glaucoma disease registry and a population-based cohort. Two replication cohorts comprised 70 JOAG and 31 PCG probands with 230 controls (MEE), and 9 PCG and 120 POAG cases (GOGS).

**Methods:** One index case per family was analysed. PRS were calculated from weighted SNPs and adjusted for ancestry. Cases were stratified by age at diagnosis (PCG ≤3 years, JOAG ≥4 and <40 years, POAG ≥40 years) and presence of pathogenic/likely pathogenic variants (“Mendelian”). Groups were compared by Kruskal-Wallis test, with logistic regression and area under the curve for per-SD effects.

**Main Outcome Measures:** PRS centile values compared across glaucoma subtypes and Mendelian and non-Mendelian subgroups.

**Results:** Mean PRS was highest in cases diagnosed in the fourth decade of life. Among subgroups, it was highest in non-Mendelian JOAG (84.1%), exceeding controls (53.3%), Mendelian JOAG (68.9%), and non-Mendelian POAG (79.3%) (all P ≤ 0.003). Non-Mendelian JOAG was also the best discriminated from controls (OR 3.66 per SD increase, 95% CI 3.02-4.49; AUC 0.814, 95% CI 0.782-0.845). Neither PCG subgroup differed from controls. Among Mendelian cases, only *MYOC* p.Gln368Ter showed significantly elevated PRS (OR 2.49 per SD, 95% CI 1.89-3.34). In the MEE replication cohort, JOAG was associated with higher PRS (OR 2.12 per SD, 95% CI 1.50-3.07) while PCG was not (OR 0.90, P = 0.73). In GOGS, mean PRS was higher in POAG than PCG (84.8 vs 59.7 centile; P = 0.026).

**Conclusions:** A POAG PRS predicts JOAG risk with discrimination comparable to or greater than POAG itself, supporting a shared polygenic architecture across juvenile and adult open-angle glaucoma. PCG showed no PRS elevation regardless of Mendelian status, indicating a distinct architecture for which rare-variant sequencing remains appropriate.

---

Open-angle glaucoma is often categorised by the age at which it is diagnosed ^1,2^. Primary open-angle glaucoma (POAG) is the latest and commonest form, with primary congenital glaucoma (PCG) representing a rarer, earlier-onset and more severe form. Between these two extremes lies juvenile open-angle glaucoma (JOAG).

Current thinking frames PCG and JOAG (collectively termed early-onset glaucoma or EOG), and POAG, as having distinct genetic architectures. In this context EOG is thought to be mostly driven by rare variants of large effect size (monogenic) ^2–7^, with POAG primarily driven by many common variants of small effect size (polygenic) ^8^. However, rare and common variants also interact, as illustrated in POAG by individuals carrying the *MYOC* p.Gln368Ter rare variant, where glaucoma penetrance and age at diagnosis are strongly influenced by polygenic risk ^9,10^.

Here we set out to test whether PCG and JOAG are polygenically continuous with POAG using a multi-trait POAG polygenic risk score (PRS). PRS are being evaluated in the clinic for stratifying risk across a range of common complex diseases, and are particularly well-suited to glaucoma given its high heritability, asymptomatic early course, and range of effective interventions ^9,11–15^. However, these scores have not been formally evaluated in the context of PCG and JOAG, which are under-ascertained or excluded from the cohorts and biobanks used for glaucoma PRS training and validation ^9^.

## Methods

### Primary study participants

Glaucoma patients were recruited through the Australian and New Zealand Registry of Advanced Glaucoma (ANZRAG) ^16^, and controls through the Genetic Risk Assessment of Degenerative Eye Disease (GRADE) study ^17^; GRADE participants were aged 50 years or older at recruitment. Written informed consent was provided under protocols approved by the Southern Adelaide Clinical Human Research Ethics Committee and adhering to the tenets of the revised Declaration of Helsinki.

Analyses were restricted to individuals of European genetic ancestry, defined as an estimated European ancestry proportion of ≥90% by principal component analysis. All secondary forms of glaucoma were excluded (pseudoexfoliative, pigmentary, angle-closure, anterior segment dysgenesis, steroid responders, and nanophthalmos). Open-angle glaucoma diagnoses were established by the referring clinician, and cases stratified by clinician-reported age at diagnosis: PCG (age at diagnosis ≤3 years), JOAG (≥4 and <40 years), and POAG (≥40 years) ^1,2^. Participants were classified as having a monogenic basis (“Mendelian”) if one or more pathogenic or likely pathogenic variants had previously been identified, noting that some monogenic variants are likely yet to be discovered or classified as disease-causing. Genes assessed included *MYOC, CYP1B1, OPTN, TBK1, TEK, THBS1*, and *ANGPT1*; cases carrying the most common *MYOC* variant (p.Gln368Ter) were additionally stratified from those carrying other pathogenic or likely pathogenic *MYOC* variants. Cases with variants in genes associated with anterior segment dysgenesis or other non-primary-open-angle glaucoma phenotypes (e.g. *FOXC1, PITX2, COL2A1, PAX6, ADAMTSL4, CPAMD8, LMX1B, NF1*) were excluded. Cases were further restricted to a single index case per family, defined as the individual with the youngest clinician-reported age at glaucoma diagnosis. Of 3,443 cases meeting all other inclusion criteria, 3,102 unrelated index cases were retained for analysis.

### Polygenic risk scores

A POAG polygenic risk score, and endophenotype scores for intraocular pressure (IOP) and vertical cup-to-disc ratio (VCDR), were developed using the multi-trait analysis of GWAS (MTAG) method as previously described (referred to in this study as “v2”) ^18^. Each score comprised approximately 7 million SNPs with non-zero weights and was adjusted for principal components of ancestry before validation. None of the ANZRAG or GRADE participants were included in the GWAS training data for these scores. An earlier version of a POAG PRS (“v1”) has been described previously ^9^ and is available via the PGS Catalog (PGS000137). The v2 scores are available as described elsewhere ^18^.

### Replication cohorts

One independent replication cohort was drawn from Mass Eye and Ear (MEE), comprising 70 JOAG probands (age at diagnosis ≥4 and <40 years), 31 PCG probands, and 230 unaffected controls who underwent whole-genome sequencing (WGS). A POAG PRS was calculated using variant weights from a cross-ancestry POAG GWAS meta-analysis ^19^ with Lassosum penalised regression trained in the UK Biobank, yielding 144,009 SNPs with non-zero weights. PRS values were normalised relative to the internal study cohort and expressed as centiles. Pairwise differences in mean PRS between groups were assessed using two-sample t-tests. Associations were assessed using multivariable logistic regression, adjusting for age, sex, genetically inferred ancestry, and variant call count.

Participants were classified as having a Mendelian basis if one or more pathogenic or likely pathogenic variants had been identified by WGS. Written informed consent was obtained under Mass General Brigham Institutional Review Board approval.

A second independent replication cohort was drawn from the Genetics of Glaucoma Study (GOGS); samples were genotyped on the Illumina Global Screening Array, with other details as described previously ^20^. We selected 9 participants with PCG (age at diagnosis ≤3 years) and 120 with POAG (age at diagnosis >40 years); all participants met the same European ancestry criteria as the discovery cohort, and the same PRS (v2 POAG) was computed. The GOGS study was approved by the QIMR Berghofer Human Research Ethics Committee, and participants provided written informed consent.

### Statistical tests

Kruskal-Wallis rank sum test was used to compare PRS centile values across all subgroups, with Dunn’s post-hoc pairwise comparisons and Benjamini-Hochberg P value adjustment. Logistic regression was used to estimate the odds ratio (OR) per standard deviation (SD) increase in PRS for each glaucoma subgroup versus controls, with the PRS standardised to the control population. Discrimination was assessed using the area under the receiver operating characteristic curve (AUC). All statistical analyses were performed in R (version 4.5.2) and figures generated using ggplot2. Logistic regressions were unadjusted since PRS values were pre-adjusted for ancestry principal components, and subgroup definitions already stratify by age.

## Results

Of 3,102 unrelated open-angle glaucoma index cases of European genetic ancestry meeting inclusion criteria, 266 were excluded from age-stratified analyses due to missing age at diagnosis, leaving 2,836 index cases. PRS values were normalised to the 1000 Genomes Project European (EUR) reference population and expressed as centiles ^18^.

Comparing clinician-reported age at glaucoma diagnosis to PRS centile (Figure 1A), we observed that the peak mean PRS value occurred in cases diagnosed in the fourth decade of life (i.e. 30-39 years). PRS values declined on either side of this peak, approaching the population mean (50th centile) in the youngest cases and decreasing more gradually in older cases.

**Figure 1.**
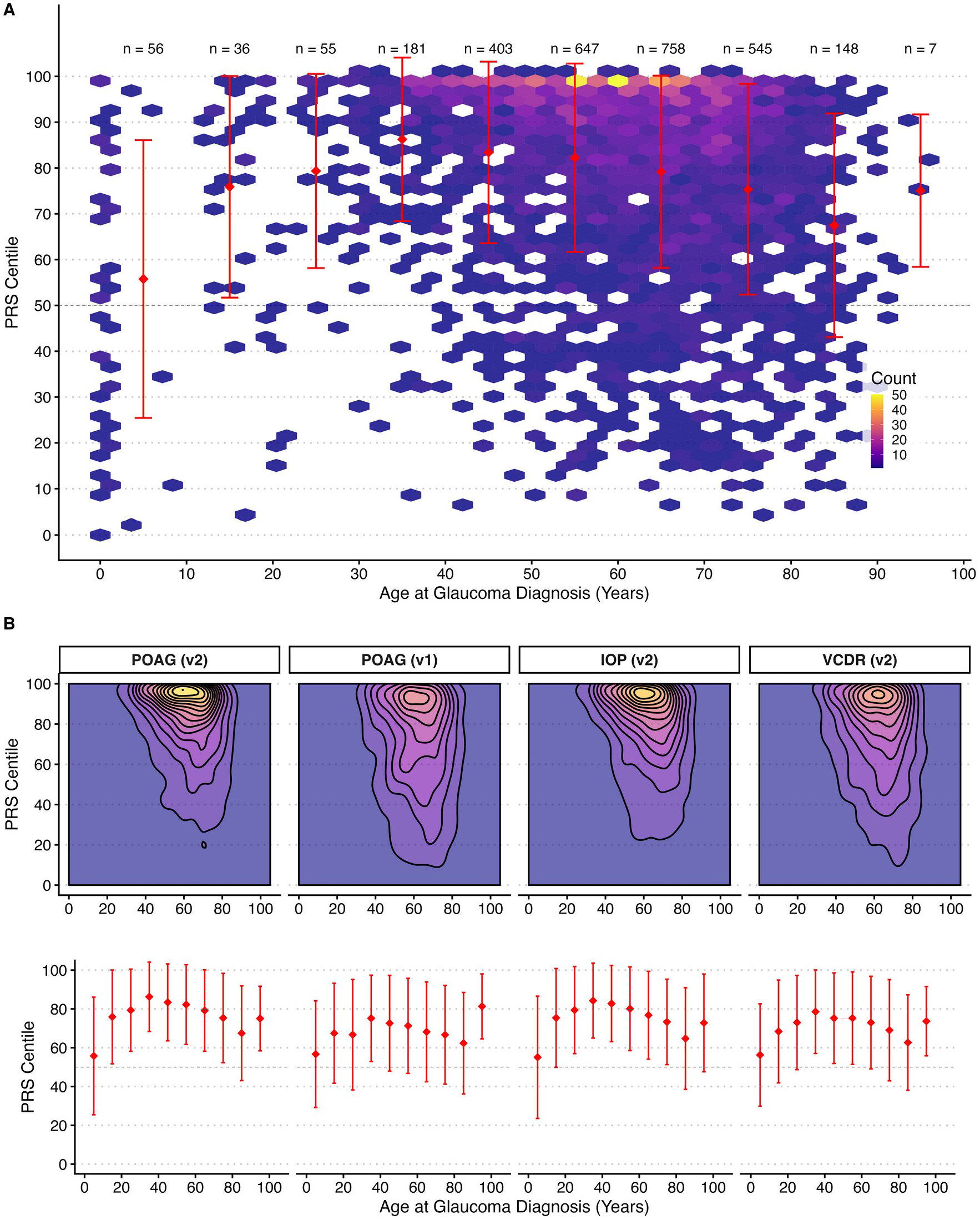
Distribution of PRS centiles as a function of age at open-angle glaucoma diagnosis. (A) Hexagonal density plot showing the distribution of ancestrally normalised PRS centiles (y-axis) against clinician-reported age at glaucoma diagnosis (x-axis) for 2,836 open-angle glaucoma index cases of European genetic ancestry. Red symbols and error bars represent mean +/− standard deviation of PRS centiles per decade. The dashed horizontal line indicates the 50th population centile. Numbers above the plot indicate case counts per decade bin. The PRS applied is version 2 of a multi-trait POAG PRS (v2 POAG ^18^). (B) Contour density plots (top) and mean +/− standard deviation per decade (bottom) comparing four polygenic scores across the same cohort: v2 POAG ^18^, v1 POAG ^9^, and endophenotype scores for intraocular pressure (v2 IOP) and vertical cup-to-disc ratio (v2 VCDR) ^18^.

We next compared the relative performance of polygenic scores tailored to predict glaucoma and its two major endophenotypes: IOP and VCDR (Figure 1B). Both endophenotype scores showed a similar age-related distribution to the POAG score, with a peak mean score for diagnoses made in the fourth decade of life (30-39 years), and a concentration of cases within the top score decile and sixth to seventh decades of life. The same pattern was seen for an earlier version of the POAG PRS (v1) ^9^, although with lower mean scores than the more recent score.

We then stratified glaucoma cases based on their clinical diagnosis of PCG, JOAG, and POAG, and by whether a Mendelian cause had been identified (Figure 2A). As previously described ^8^, the ratio of Mendelian to non-Mendelian forms of disease was higher in PCG (~1:3) and JOAG (~1:8) than for POAG (~1:43), representing diagnostic rates of 25%, 11%, and 2% respectively (Table 1). All glaucoma cases were compared to an ancestrally matched control cohort recruited from the same populations ^17^.

**Table 1.** Baseline characteristics of the study cohort. PCG is defined by an age at diagnosis of ≤3 years, JOAG as ≥4 and <40 years, and POAG as ≥40 years. Mendelian refers to cases with pathogenic or likely pathogenic variants identified in a Mendelian pattern of inheritance. Demographic data were available for only a small subset of controls and are not shown (—).

|  | Contr<br>ol | PCG<br>Mendelia<br>n | PCG non-Mendelian | JOAG<br>Mendelian | JOAG non-Mendelian | POAG<br>Mendelian | POAG non-Mendelian |
| --- | --- | --- | --- | --- | --- | --- | --- |
| N | 999 | 13 | 39 | 24 | 202 | 58 | 2500 |
| Age at<br>diagnosis, years<br>(median (IQR)) | — | 0 (0-0) | 0 (0-0) | 30 (21-34) | 34 (26-37) | 55 (49-65) | 63 (55-72) |
| Female sex (n<br>(%)) | — | 8 (62%) | 16 (41%) | 11 (46%) | 96 (48%) | 27 (47%) | 1384 (55%) |
| PRS centile<br>(mean (SD)) | 53.3<br>(29.7) | 49.9 (32.2) | 57.5 (29.0) | 68.9 (25.2) | 84.1 (20.0) | 75.3 (23.2) | 79.3 (21.6) |

**Figure 2.**
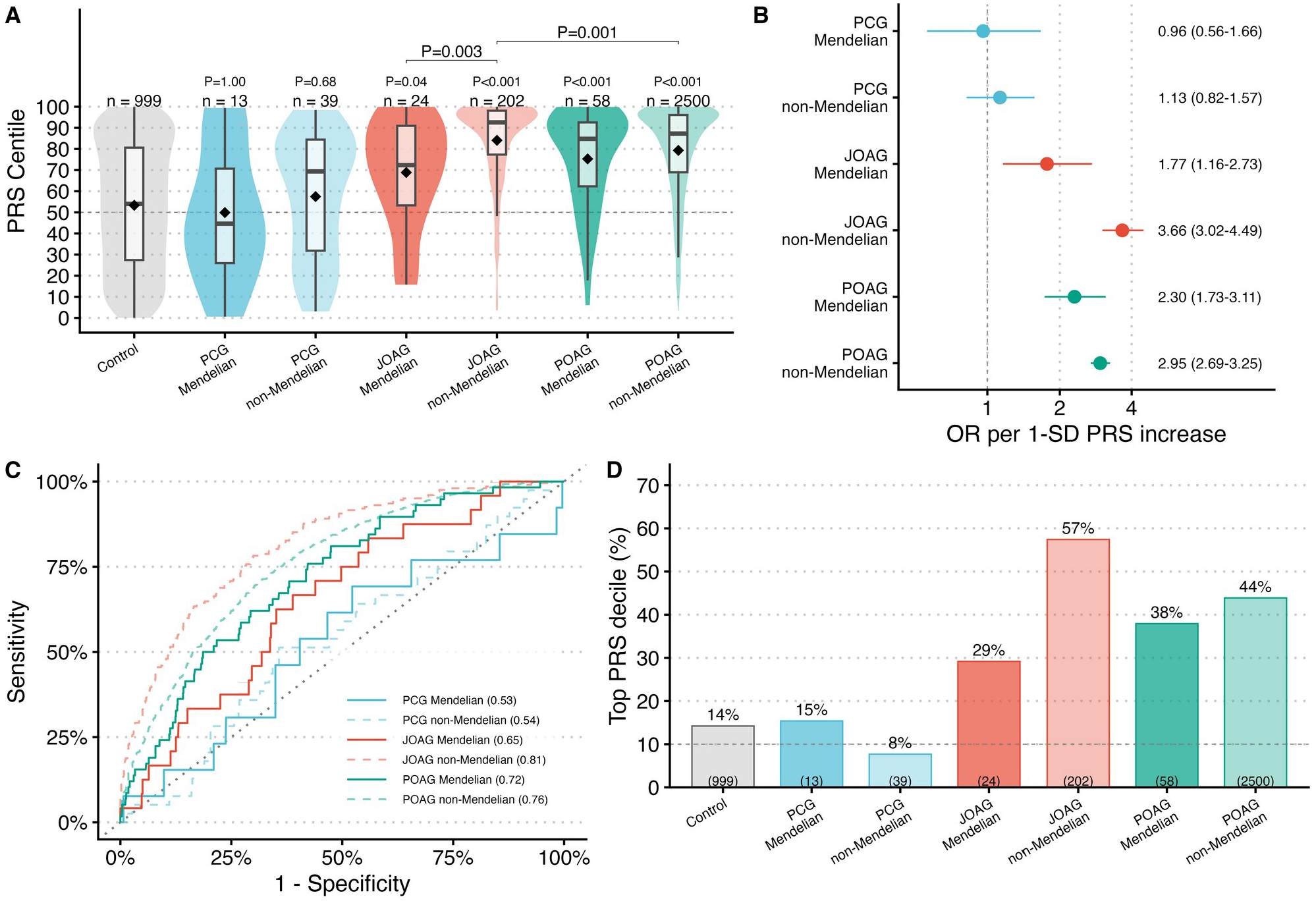
Comparisons of glaucoma polygenic risk scores between glaucoma subtypes. (A) Violin and box plots showing the mean (black diamond), median (horizontal line), interquartile range (box), 1.5 × IQR (whiskers), and outliers of PRS centiles across glaucoma subtypes and controls. The dashed horizontal line indicates the 50th population centile. Numbers above each box indicate subgroup sizes. Benjamini-Hochberg-adjusted P values from Dunn’s post-hoc comparisons against controls are shown above each group, and above the brackets for two selected between-subgroup comparisons (JOAG non-Mendelian vs POAG non-Mendelian, and vs JOAG Mendelian). Kruskal-Wallis P < 2.2 × 10^−16^. Boxes are colour-coded by diagnosis: blue (PCG), red (JOAG), teal (POAG), grey (controls); darker shading indicates Mendelian cases and lighter shading indicates non-Mendelian cases. (B) Forest plot of odds ratios (OR) per standard deviation (SD) PRS increase for each subgroup vs. controls, with 95% confidence intervals on a log scale. The dashed vertical line indicates an OR of 1 (no effect). (C) Receiver operating characteristic curves for each subgroup vs. controls, with AUC values in parentheses. (D) Proportion of each subgroup falling within the top population PRS decile (>=90th centile). The dashed line indicates the expected 10% under a null distribution. Numbers in parentheses indicate subgroup sizes.

The mean PRS centile among controls was 53.3%, marginally above the 50th population centile (Table 1, Figure 2A). The highest mean PRS centile was observed in non-Mendelian JOAG cases (84.1%), consistent with observations made in Figure 1A. This was significantly higher than both non-Mendelian POAG (79.3%) and Mendelian JOAG (68.9%) (Dunn’s test, adjusted P = 0.001 and P = 0.003 respectively). These findings suggest that a POAG-trained PRS will have utility in JOAG as well as POAG, and that JOAG and POAG likely represent continua of the same disease pathophysiology.

Both JOAG and POAG had lower mean PRS values in their Mendelian form (68.9%, 75.3% respectively) compared to their non-Mendelian forms (84.1%, 79.3% respectively) (Table 1; Supplementary Table S1). Nonetheless, these were both still higher than controls (Dunn’s adjusted P = 0.04 and P < 0.001 respectively; Figure 2A), suggesting that polygenic risk can still be additive in the context of Mendelian variants, as previously described for the incomplete penetrance variant *MYOC* p.Gln368Ter in POAG ^9^, which is the most common variant in the POAG Mendelian group presented here.

Unlike JOAG and POAG, the mean PRS in PCG did not differ from controls (Table 1, Figure 2A). This was true for both Mendelian and non-Mendelian forms (P = 1.00 and 0.68 respectively), suggesting that unlike JOAG, PCG is a disease in which POAG-associated common variants do not appear to play a major role. In an unadjusted logistic regression, each SD increase in PRS was associated with an odds ratio of 3.66 (95% CI 3.02-4.49; P < 0.001) for non-Mendelian JOAG vs. controls, 2.95 (95% CI 2.69-3.25; P < 0.001) for non-Mendelian POAG vs. controls, and 1.13 (95% CI 0.82-1.57; P = 0.46) for non-Mendelian PCG vs. controls (Figure 2B). Mendelian subgroups showed a similar pattern: OR 1.77 (95% CI 1.16-2.73; P = 0.009) for Mendelian JOAG and 2.30 (95% CI 1.73-3.11; P < 0.001) for Mendelian POAG, while Mendelian PCG did not differ from controls (OR 0.96; 95% CI 0.56-1.66; P = 0.88). The AUC for discriminating non-Mendelian JOAG from controls was 0.814 (95% CI 0.782-0.845), compared to 0.761 (95% CI 0.744-0.779) for non-Mendelian POAG and 0.538 (95% CI 0.448-0.628) for non-Mendelian PCG (Figure 2C). The enrichment of cases in the top PRS decile further illustrates this pattern: 57% of non-Mendelian JOAG cases fell in the top population decile, compared to 44% of non-Mendelian POAG, 8% of non-Mendelian PCG, and 14% of controls (Figure 2D). Odds ratios and AUCs across all four polygenic scores are provided in Supplementary Table S2.

In an independent replication cohort (MEE), JOAG was independently associated with a higher PRS in multivariable analysis (OR 2.12 per SD increase, 95% CI 1.50-3.07; P = 3.38 × 10^−5^), while PCG was not (OR 0.90, 95% CI 0.50-1.68; P = 0.73) (Figure 3A). In unadjusted pairwise comparisons (two-sample t-tests), PRS was significantly higher in JOAG than in controls (P = 0.0017) and than in PCG (P = 0.021), whereas PCG did not differ from controls (P = 0.54). Non-Mendelian JOAG cases had higher PRS compared to Mendelian JOAG (mean centile 58.5 vs 44.7), although this difference did not reach statistical significance (P = 0.10; Figure 3B). In PCG, PRS centiles did not differ between Mendelian and non-Mendelian cases (mean centile 41.7 vs 36.1; P = 0.62; Figure 3B). In a second independent replication cohort (GOGS), mean PRS was significantly higher in POAG than in PCG (84.8% vs 59.7%; P = 0.026, two-sample t-test).

**Figure 3.**
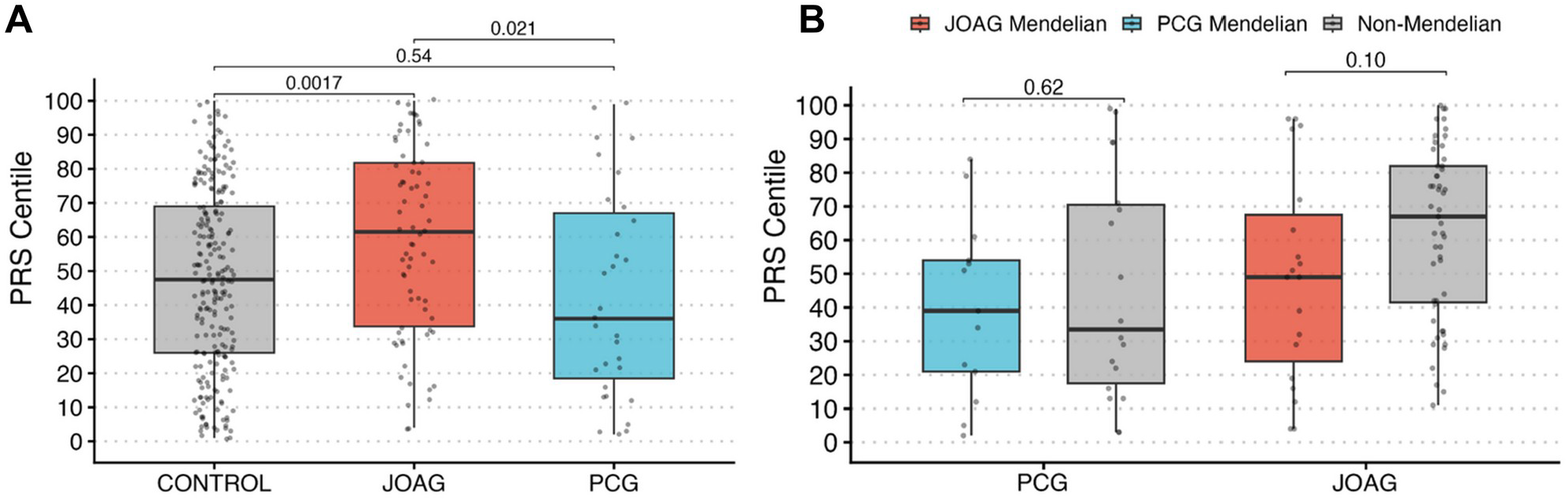
Independent replication of PRS associations. (A) Box plots showing PRS distribution in centiles across controls, PCG, and JOAG in an independent cohort of 331 individuals. P values shown are from unadjusted pairwise two-sample t-tests; the multivariable logistic regression results reported in the text are adjusted for age, sex, ancestry, and variant call count. (B) PRS centiles in JOAG and PCG grouped by Mendelian and non-Mendelian status. Sample sizes: Mendelian PCG (n=13), non-Mendelian PCG (n=18), Mendelian JOAG (n=19), non-Mendelian JOAG (n=51).

Finally, we examined the PRS distribution among Mendelian cases stratified by causal gene (Figure 4). Gene groups differed in their age-at-onset composition: biallelic *CYP1B1* variants were predominantly associated with congenital-onset glaucoma, whereas *MYOC* p.Gln368Ter was predominantly associated with adult-onset glaucoma (Figure 4A). Among individuals carrying *MYOC* variants, those with the common p.Gln368Ter variant had a significantly higher mean PRS than controls (OR 2.49 per SD increase, 95% CI 1.89-3.34; P < 0.001; Figure 4B, 4C), while those carrying other *MYOC* variants did not (OR 1.26, 95% CI 0.67-2.40). Individuals carrying variants in other glaucoma-associated Mendelian genes (*OPTN, TBK1*, and others) also showed no significant PRS elevation (OR 1.17, 95% CI 0.66-2.10), and individuals with biallelic *CYP1B1* variants, who typically present with congenital rather than open-angle glaucoma, had PRS values indistinguishable from controls (OR 0.85, 95% CI 0.43-1.70). Those carrying *MYOC* p.Gln368Ter had a mean PRS centile (77) comparable to non-Mendelian POAG (79; Figure 4D), and enrichment in the top population PRS decile was most pronounced for this group (40%, vs 20% for other *MYOC* variants, 17% for other Mendelian genes, and 12% for biallelic *CYP1B1*; Figure 4E). Mendelian POAG was almost entirely attributable to *MYOC* (55/58, 95%), predominantly p.Gln368Ter (54/58, 93%). These findings remain consistent with *MYOC* p.Gln368Ter acting as a variant of incomplete penetrance whose clinical expression is modulated by polygenic background ^9,10^.

**Figure 4.**
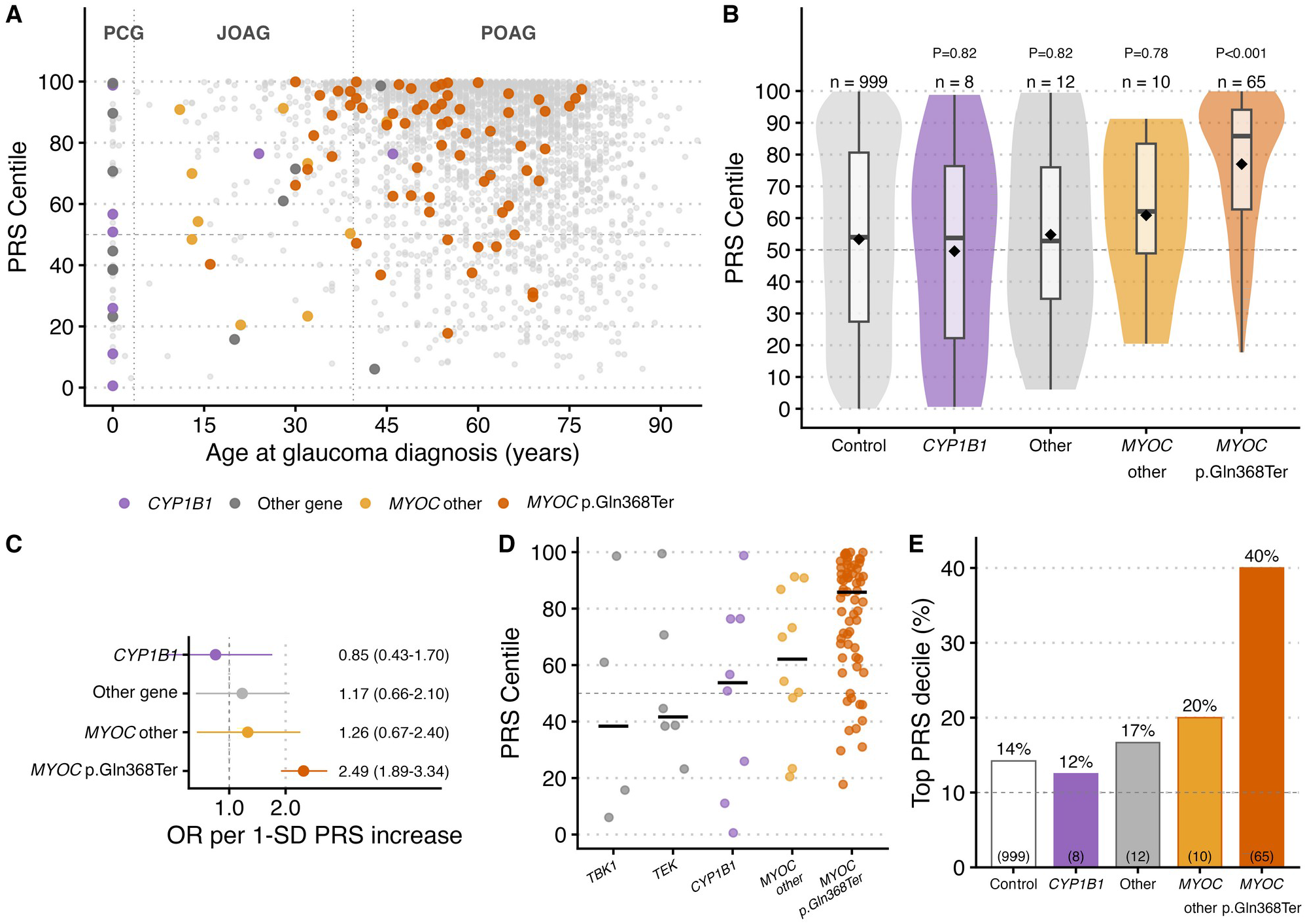
Polygenic risk score by age at diagnosis and causal gene across glaucoma subtypes. (A) PRS centile versus age at glaucoma diagnosis for all glaucoma cases (grey), with Mendelian cases highlighted by causal gene. Dotted vertical lines mark the boundaries between PCG, JOAG, and POAG; the dashed line marks the 50th PRS centile. (B) Violin and box plots comparing PRS centiles between controls and Mendelian cases stratified by causal gene: *CYP1B1*, other Mendelian glaucoma genes (*OPTN, TBK1, TEK, ANGPT1*), other *MYOC* variants, and *MYOC* p.Gln368Ter. Formatting as per Figure 2A. (C) Forest plot of odds ratios per SD PRS increase for each gene group versus controls, with 95% confidence intervals on a log scale. (D) Individual-level PRS centiles for Mendelian glaucoma cases, grouped by causal gene (genes with three or more cases); crossbar indicates median. (E) Proportion of each gene group falling within the top population PRS decile (≥90th centile). The dashed line indicates the expected 10% under a null distribution.

## Discussion

Counter to the prevailing view that EOG is driven primarily by rare variants of large effect size ^2–7^, we find that non-Mendelian JOAG carries the highest mean polygenic burden of any subgroup in this cohort, higher than adult-onset POAG itself. Indeed, a POAG-trained PRS had better discriminating power for JOAG than POAG (AUC against controls of 0.81 vs 0.76), and placed 57% of non-Mendelian JOAG cases in the top population decile.

These findings reframe JOAG and POAG as a single polygenic continuum rather than disease with distinct genomic architectures, with JOAG representing the upper tail of shared common-variant risk combined with additional rare-variant burden in a minority of cases. The age-related boundary between JOAG and POAG (at 40 years) should therefore not be considered a pathophysiological divide.

Our findings predict that POAG PRS will also have utility in JOAG, particularly for the majority (~89%) who do not have a known Mendelian genetic basis. Siblings of JOAG patients who carry a high PRS may benefit from earlier and more frequent glaucoma screening, even in the absence of known Mendelian causes, while PRS may also help guide treatment intensity in early disease, as reported in POAG ^13–15^.

Even in the presence of a Mendelian variant, the elevation of mean PRS in JOAG suggests that polygenic risk modulates the penetrance of these variants, as described previously in POAG and a range of other common complex diseases ^9,21,22^. Our gene-specific analysis reinforces this point: individuals carrying *MYOC* p.Gln368Ter, a variant of well-established incomplete age-related penetrance ^9,10^, showed a significantly elevated PRS relative to controls, consistent with polygenic risk modulating the disease onset associated with this variant.

In contrast to JOAG, neither this PRS nor scores tuned to predict IOP or VCDR were informative for PCG risk; current glaucoma PRS tests are therefore not likely to be informative in PCG, and genetic evaluation in these families should instead prioritise rare variant testing. It remains possible that a different subset of common variants can modulate PCG risk, although assembling cohorts large enough to detect these will remain a significant challenge.

As proposed in other conditions, a below-median PRS (below the 50th population centile) may raise the prior probability that patients diagnosed with JOAG or POAG carry rare variants with Mendelian inheritance. However, since 79% of our Mendelian JOAG cases had an above-median PRS (69% after excluding *MYOC* p.Gln368Ter), rare variant testing therefore remains warranted across JOAG irrespective of PRS. Current technologies require different testing for monogenic and polygenic causes, but as sequencing costs decline, combined rare- and common-variant testing may become a more cost-effective option.

To our knowledge, this study is the first to investigate the role of a PRS across the complete open-angle glaucoma age spectrum, benefiting from large numbers of carefully phenotyped cases that are under-ascertained or excluded from population biobanks.

Limitations include small PCG subgroups (n=13 Mendelian, n=39 non-Mendelian), meaning that modest PRS elevations cannot be excluded for PCG. Controls from the GRADE study were aged 50 years and older, creating an age asymmetry with PCG and JOAG cases, although PRS centiles were normalised to an ancestrally matched population. Cases and controls were ascertained separately and therefore reported AUC values likely overestimate performance in unselected or population-screening settings. It is also likely that other Mendelian variants remain to be found, which means these values are likely an underestimate of the true difference in PRS values between Mendelian and non-Mendelian groups. These findings were independently replicated in two cohorts: the first, a using a methodologically distinct POAG PRS derived from cross-ancestry GWAS data ^19^, confirmed PRS elevation in JOAG but not PCG, and showed a non-significant trend towards higher PRS in non-Mendelian than Mendelian JOAG cases (Figure 3); the second, using the same PRS as our primary cohort, showed despite small sample sizes that PCG differs from adult-onset POAG in PRS distribution. These findings have not yet been validated in non-European populations, although this may soon be possible with the emergence of larger and more ancestrally diverse cohorts ^23^.

## Abbreviations

AUC: area under the receiver operating characteristic curve
EOG: early-onset glaucoma
GOGS: Genetics of Glaucoma Study
IOP: intraocular pressure
JOAG: juvenile open-angle glaucoma
MEE: Mass Eye and Ear
MTAG: multi-trait analysis of genome-wide association studies
PCG: primary congenital glaucoma
POAG: primary open-angle glaucoma
PRS: polygenic risk score
SNP: single nucleotide polymorphism
VCDR: vertical cup-to-disc ratio
WES: whole-exome sequencing
WGS: whole-genome sequencing

## Institutional Review Board

Institutional Review Board (IRB)/Ethics Committee approval was obtained (Southern Adelaide Clinical Human Research Ethics Committee). MGB (Mass General Brigham) Institutional Review Board approval was obtained for the Mass Eye and Ear replication cohort, and the Genetics of Glaucoma Study was approved by the QIMR Berghofer Human Research Ethics Committee.

## Data Availability

Polygenic score weights are available through the PGS Catalog (PGS000137, version 1 POAG) and as previously described^18^ (version 2 scores). Individual-level cohort data are not publicly available owing to participant privacy and consent restrictions.

## Supplementary Material

**Supplementary Table S1.** Polygenic risk score centile distribution across glaucoma subgroups. Number of individuals (N) and the mean, median, and standard deviation (SD) of PRS centile in each subgroup and in controls. PRS centiles were normalised to the 1000 Genomes Project European reference population.

| Group | N | Mean | Median | SD |
| --- | --- | --- | --- | --- |
| Control | 999 | 53.3 | 54.0 | 29.7 |
| PCG Mendelian | 13 | 49.9 | 44.6 | 32.2 |
| PCG non-Mendelian | 39 | 57.5 | 69.4 | 29.0 |
| JOAG Mendelian | 24 | 68.9 | 72.3 | 25.2 |
| JOAG non-Mendelian | 202 | 84.1 | 92.6 | 20.0 |
| POAG Mendelian | 58 | 75.3 | 84.8 | 23.2 |
| POAG non-Mendelian | 2500 | 79.3 | 87.3 | 21.6 |

**Supplementary Table S2.** Odds ratios and discrimination across four polygenic scores. Odds ratio (OR) per standard deviation increase in PRS and area under the receiver operating characteristic curve (AUC), each with 95% confidence intervals, for each glaucoma subgroup versus controls, across four polygenic scores: the version 2 multi-trait POAG score, the version 1 POAG score, and endophenotype scores for intraocular pressure (IOP) and vertical cup-to-disc ratio (VCDR). The JOAG non-Mendelian subgroup (in bold) had the highest OR and AUC for all four scores.

| Subgroup | OR (POAG (v2)) | AUC (POAG (v2)) | OR (POAG (v1)) | AUC (POAG (v1)) | OR (IOP (v2)) | AUC (IOP (v2)) | OR (VCDR (v2)) | AUC (VCDR (v2)) |
| --- | --- | --- | --- | --- | --- | --- | --- | --- |
| PCG Mendelian | 0.96 (0.56-1.66) | 0.529 (0.353-0.704) | 0.90 (0.52-1.55) | 0.509 (0.370-0.647) | 0.76 (0.44-1.31) | 0.554 (0.374-0.734) | 1.14 (0.66-2.00) | 0.503 (0.329-0.676) |
| PCG non-Mendelian | 1.13 (0.82-1.57) | 0.538 (0.448-0.628) | 1.30 (0.95-1.80) | 0.573 (0.477-0.668) | 1.18 (0.86-1.63) | 0.554 (0.455-0.652) | 1.14 (0.83-1.59) | 0.541 (0.461-0.621) |
| JOAG Mendelian | 1.77 (1.16-2.73) | 0.652 (0.549-0.754) | 1.59 (1.06-2.39) | 0.637 (0.529-0.744) | 1.81 (1.21-2.71) | 0.652 (0.530-0.773) | 1.29 (0.85-1.96) | 0.570 (0.473-0.667) |
| **JOAG non-Mendelian** | 3.66 (3.02-4.49) | 0.814 (0.782-0.845) | 2.31 (1.96-2.73) | 0.721 (0.683-0.759) | 3.47 (2.89-4.22) | 0.812 (0.780-0.845) | 2.85 (2.39-3.44) | 0.761 (0.726-0.797) |
| POAG Mendelian | 2.30 (1.73-3.11) | 0.718 (0.655-0.781) | 2.18 (1.66-2.90) | 0.713 (0.646-0.780) | 2.55 (1.92-3.42) | 0.749 (0.683-0.814) | 1.60 (1.22-2.11) | 0.625 (0.549-0.700) |
| POAG non-Mendelian | 2.95 (2.69-3.25) | 0.761 (0.744-0.779) | 2.00 (1.85-2.18) | 0.681 (0.662-0.701) | 2.73 (2.50-2.99) | 0.752 (0.734-0.770) | 2.24 (2.06-2.44) | 0.707 (0.688-0.726) |

